# Premorbid predictors of death at initial presentation of coronary heart disease in the Women’s Health Initiative study

**DOI:** 10.1101/2024.04.12.24305749

**Authors:** Ming-Li Chen, Jin Li, Kruthika R. Iyer, Catherine Tcheandjieu, Shirin Jimenez, Elias Levy Itshak Salfati, Liana C. Del Gobbo, Marcia L Stefanick, Manisha Desai, Xiaonan Xue, Themistocles L Assimes

**Author notes:** **Address for correspondence:** Themistocles L. Assimes, MD PhD Palo Alto VA Hospital, 3801 Miranda Ave., Palo Alto, CA 94304.

## Abstract

**Background:** Premorbid health traits that predispose to a fatal initial presentation of coronary heart disease (CHD) remain poorly characterized.

**Methods:** We followed 148,230 post-menopausal participants in the Women’s Health Initiative for a median of 13.3 years. We ascertained the first occurrence of CHD and performed a joint Cox multivariate regression to identify premorbid predictors for a fatal rather than a non-fatal incident event.

**Results:** A total of 10,714 incident CHD events including 513 fatal events accrued during follow up. A five-year increase in age, smoking 5 to 34 cigarettes per day, and a standard deviation (SD) increase in the Cornel voltage product on electrocardiography each independently increased the relative risk (RR) of dying from one’s initial presentation of CHD by 46% (95% confidence interval [CI], 35 to 58%), 30% (8 to 51%,), and 17% (7 to 28%), respectively. A high level of recreational physical activity (>1200 metabolic equivalent (MET) minutes per week) reduced one’s relative risk by 32% (12 to 49%). A significant dose-response effect was observed for both physical activity and smoking and the reduction in absolute risk of presenting with fatal CHD associated with a healthy lifestyle was roughly equivalent to the difference in risk observed among women separated in age by approximately 10 years.

**Conclusions:** Modifiable factors affect a post-menopausal woman’s risk of dying from her initial presentation of CHD. Our findings may reduce case-fatality rates of CHD by motivating individuals at risk to adopt and/or adhere to established primary prevention strategies.

## Introduction

Initial presentations of coronary heart disease (CHD) include stable angina, acute coronary syndrome (ACS), and myocardial infarction (MI). A minority of subjects suffer a fatal consequence from their initial presentation, but this outcome is arguably one of the most devastating events encountered by patients, their family members, and healthcare providers.^1^ Individuals dying from their initial presentation of CHD miss the opportunity to benefit from well-established secondary prevention strategies that reduce the risk of recurrent events.^2^

Multiple studies have demonstrated a sustained decline in the overall rates of CHD over the last few decades in developed countries.^3^ This decline comes on the heels of the implementation of several primary and secondary prevention strategies.^2,4^ However, the decline in the rates of fatal CHD has not been as steep as CHD overall.^1,5^ Some studies have shown that the rate of fatal CHD has remained stable over time among subjects with no prior history of CHD and may have even increased among elderly individuals.^5^ Furthermore, lifetime risks of CHD and sudden cardiac death in the community remain high.^6,7^ These trends underscore the need to identify premorbid predictors of a fatal outcome related to incident CHD.

The premorbid predictors for a fatal outcome with the initial presentation of CHD remain poorly characterized. In the Cardiovascular Health Study (CHS), the only independent predictors of fatal CHD outcomes among subjects with no prior history of CHD, MI, or CHF at baseline were diabetes and age^8^. The study was limited by the small number of events, the narrow range of age of elderly participants, and the lack of assessment of lifestyle factors. In this study, we leveraged the large size and the prolonged follow-up of the Women’s Health Initiative (WHI) in combination with the repeated assessment of exposures and the careful adjudication of CHD outcomes to document independent predictors of a fatal presentation of CHD more reliably.

## Methods

### Study cohort

A detailed description of study design, recruitment, and initial characterization for WHI has been previously published.^9–11^ The study cohort consisted of 148,230 participants enrolled in either one of the clinical trials (n=63170) or the observational study (n=85060) between 1993 and 1998 with no history of atherosclerotic cardiovascular disease or congestive heart failure at baseline (**Figure 1**). All participants were followed through 2010 and a subset of participants (n=88550) with continued adjudication of CHD outcomes were further followed until 2015. The WHI project was reviewed and approved by the Fred Hutchinson Cancer Research Center (Fred Hutch) Institutional Review Board (IRB). Participants provided written informed consent to participate. Additional consent to review medical records was obtained through signed written consent.

**Figure 1.**
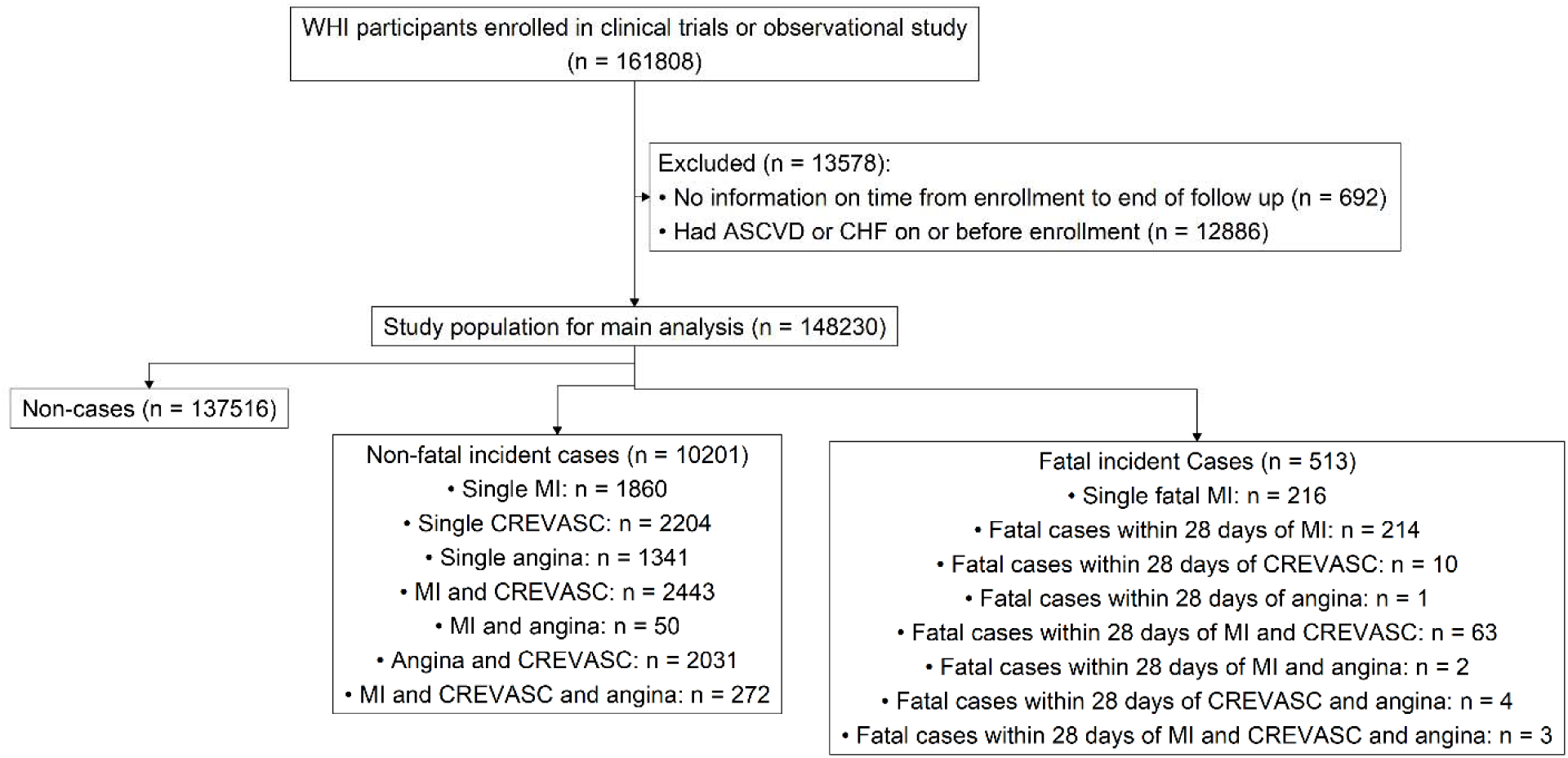
Flow diagram of Women’s Health Initiative participants and outcomes examined in this study during the period 1993-2015. ASCVD: atherosclerosis related cardiovascular disease, CHF: congestive heart failure, CHD: coronary heart disease; CHD cases include individuals whose initial presentation of CHD during follow up was adjudicated as myocardial infarction (MI), a coronary revascularization procedure (CREVASC), a definite death from CHD. CHD cases are considered fatal if the participant died within 28 days of her first CHD event.

### Outcome ascertainment

The primary endpoints for this study were newly diagnosed CHD events including angina, non-fatal myocardial infarction (MI), coronary revascularization through either a percutaneous coronary intervention or coronary artery bypass grafting, and definitive death from CHD. Adjudication methods for cardiovascular disease outcomes in the WHI have been previously described extensively^12^. The diagnosis of nonfatal MI was confirmed if data in the hospital record met standardized criteria of diagnostic electrocardiographic changes, elevated cardiac enzyme levels, or both. Treatment with coronary revascularization was confirmed by documentation of the procedure in the medical record. Fatal CHD was considered confirmed if there was documentation in the hospital or autopsy records or if coronary disease was listed as the cause of death on the death certificate and evidence of previous coronary disease was available. Adjudication of these events occurred for all participants through the clinical trial and intervention phase (1993-2005) as well as the first extension (2005-2010). Adjudication of CHD events during the second extension study (2010-2015) was restricted to a subset of participants in the Medical Record Cohort (MRC). We considered all adjudicated events occurring within 28 days of the initial event in classifying whether the initial presentation was fatal or not fatal. Thus, women who had a fatal event within 28 days of their initial non-fatal event were classified as fatal events.

### Exposure Assessment

Information on exposures was collected at baseline through an in-person clinic visit and through multiple self-administered questionnaires that collected information on demographic factors including age, race/ethnicity, education, income, medical history, physical activity (PA) levels, smoking habits, and diet. Women who participated in any clinical trial also underwent a 12-lead electrocardiogram (ECG). Additional self-administered questionnaires were collected regularly during follow up allowing for repeated measures of medical history, medication use, PA levels, and smoking habits. The data collection schedule during follow-up varied by cohort (clinical trials vs. observational study) and by the study phase (intervention vs. observational phase). Complete details on the collection schedule and the derivation of variables are provided in **Table S1**.

We used the medication inventory to document the use of aspirin, statins, beta-blockers, calcium channel blockers (CCBs), inhibitors of the renin-angiotensin system (RAS), and diuretics. Participant’s level of PA was categorized into four groups based on weekly energy expenditure in metabolic equivalents (MET) minutes.^13^ In addition to typical ECG intervals, we tested several derived ECG variables related to left ventricular mass/hypertrophy and ECG abnormalities indicating possible subclinical MI/Ischemia or bradycardia. Through six ancillary studies, a subset of all WHI participants underwent genome-wide genotyping followed by joint imputation to the 1000G reference panel (dbGaP study accession: phs000746.v1.p3) We used this data to construct a previously validated genetic risk score for CHD (metaGRS) consisting of genetic variants summarizing a participant’s exposure to high-risk variants at 1.7 million polymorphic sites^14^. We restricted the metaGRS analysis to non-Hispanic white participants given the score was constructed using almost exclusively a non-Hispanic white population and has been shown to perform best in the same population^14^. Through separate ancillary studies, seven blood-based biomarkers related to cardiovascular disease were measured using standardized assays in a subset of WHI participants including high density lipoprotein concentration (HDL), low density lipoprotein concentration (LDL), triglycerides, creatinine, insulin, glucose, and high sensitivity C-reactive protein (hsCRP). Complete details of exposure assessment are in **Supplemental Material**.

### Statistical Analyses

We used a joint multivariate Cox model to estimate the hazard ratio of a predictor of interest with each of the two subtypes of presentation of incident CHD, fatal and non-fatal, simultaneously.^15–18^ More extensive details on the statistical modeling implemented is provided in the **Supplemental Material**. Predictors of interest were included in a multivariate model if they demonstrated a significant association with CHD in a minimally adjusted model that included age and race. Among the predictors of interest, we modeled body mass index (BMI), systolic blood pressure (SBP), smoking status, amount of weekly recreational physical activity, treated hypertension, treated diabetes, and class of medication use as time-dependent covariates while the other risk factors measured at baseline were modeled as time-fixed covariates. For main analyses, missingness of covariates were imputed with multiple imputation method, using linear mixed effect where covariates were modeled mostly as random effect, resulting in a predicted value close to the predicted value of the missing sample^19,20^. We corrected for multiple testing of predictors included in the multivariate model using the Benjamini-Hochberg method and a false discovery rate (FDR) of 0.05^21^. Lastly, we further performed subgroup analyses limited to the subsets of participants with ECG measures, blood biomarkers, and genetic data. These variables were tested by adding them last to the main model containing the covariates included in the main analyses.

Next, we used the same joint multivariate Cox model to estimate the probability of i. developing incident non-fatal CHD over a follow up period of 20 years and ii. the probability of a fatal incident CHD event taking into consideration the competing risk of a non-fatal CHD event. We calculated and compared these hazard functions for women at different ages and with various combination of exposures of interest based on the results of our multivariate analysis.

All analyses were performed using R.

## Results

A total of 10201 non-fatal and 513 fatal incident CHD events accrued among 148,230 participants during a median follow-up time of 13.3 years for an overall incident CHD rate of 7.2% and a case-fatality rate of 5.0% (**Figure 1**, **Table 1**). Compared to women with no events, women with an incident CHD event during follow up were older, had a lower socioeconomic status, more often were current smokers, and had a lower degree of recreational physical activity (PA) at baseline (**Table 1**). Women with CHD events also had a higher BMI and blood pressure, as well as a higher prevalence of treated hypertension and treated diabetes. Lastly, women with CHD events also had a higher use of aspirin, statins, beta-blockers, calcium channel blockers, renin-angiotensin inhibitors, and thiazide-like diuretics (**Table 1**).

**Table 1.** Baseline characteristics of study participants.

| Characteristics | Non-event |  | Non-fatal CHD |  | Definitive fatal CHD |  |
| --- | --- | --- | --- | --- | --- | --- |
|  | (N = 137516) |  | (N = 10201) |  | (N = 513) |  |
|  | N | Mean (SD) or median [IQR] or % | N | Mean (SD) or median [IQR] or % | N | Mean (SD) or median [IQR] or % |
| **Demographics/anthropometric** |  |  |  |  |  |  |
| Age (years) | 137516 | 62.7 (7.2) | 10201 | 65.6 (6.7) | 513 | 68.6 (6.5) |
| Race (%) |  |  |  |  |  |  |
| European American | 113878 | 83.0 | 8703 | 85.5 | 440 | 85.9 |
| African American | 11833 | 8.6 | 879 | 8.6 | 50 | 9.8 |
| Hispanics | 5547 | 4.0 | 308 | 3.0 | 10 | 2.0 |
| Asians or Pacific Islander | 3794 | 2.8 | 142 | 1.4 | 6 | 1.2 |
| BMI (kg/m<sup>2</sup>) | 136322 | 27.8 (5.9) | 10123 | 29.0 (6.0) | 508 | 29.1 (6.5) |
| SBP (mmHg) | 137408 | 128.5 (19.1) | 10194 | 137.3 (19.8) | 512 | 140.2 (20.7) |
| **Medical history (%)** |  |  |  |  |  |  |
| Treated hypertension | 137516 | 21.6 | 10201 | 35.7 | 513 | 40.9 |
| Treated diabetes | 137516 | 3.1 | 10201 | 10.1 | 513 | 11.5 |
| **Lifestyles and social economic factors** |  |  |  |  |  |  |
| Current smoker (%) | 137427 | 6.5 | 10195 | 8.6 | 512 | 10.2 |
| Physical activity (MET hr/week) | 137427 | 8.0 [1.5, 17.5] | 10195 | 6.2 [0.8, 15.2] | 512 | 5.8 [0.5, 14.7] |
| AHEI score | 134880 | 59.0 (11.5) | 10000 | 57.7 (11.3) | 503 | 58.8 (10.8) |
| Education level | 136491 | 7.3 (1.9) | 10120 | 6.9 (1.9) | 508 | 7.1 (1.9) |
| Family income | 131994 | 4.3 (1.8) | 9818 | 3.8 (1.7) | 492 | 3.7 (1.8) |
| **Medication use (%)** |  |  |  |  |  |  |
| Aspirin | 137514 | 18.6 | 10201 | 25.0 | 513 | 20.9 |
| Statin | 137514 | 6.1 | 10201 | 10.3 | 513 | 9.2 |
| Beta-blocker | 137514 | 6.9 | 10201 | 11.6 | 513 | 10.7 |
| CCB | 137514 | 7.8 | 10201 | 16.1 | 513 | 17.9 |
| RAS inhibitor | 137514 | 7.6 | 10201 | 12.4 | 513 | 14.6 |
| Diuretics | 137514 | 10.6 | 10201 | 16.2 | 513 | 18.7 |
| **ECG measures** |  |  |  |  |  |  |
| Bradycardia (%) | 53499 | 2.6 | 4615 | 2.9 | 216 | 2.3 |
| LVHI | 53132 | 80.0 [76.0, 84.0] | 4584 | 82.0 [78.0, 87.0] | 212 | 82.0 [78.0, 88.0] |
| LVHI_CAT (%) | 53132 | 0.2 | 4584 | 0.7 | 212 | 3.3 |
| NOVA5 (%) | 51735 | 28.6 | 4383 | 40.2 | 197 | 44.7 |
| MC_MI (%) | 51667 | 2.0 | 4378 | 3.7 | 197 | 5.6 |
| **Biomarkers** |  |  |  |  |  |  |
| HDLC (mg/dL) | 29169 | 55.5 (14.4) | 4624 | 51.1 (13.8) | 293 | 51.5 (14.6) |
| LDLC (mg/dL) | 24127 | 143.8 (37.4) | 3838 | 148.6 (37.9) | 239 | 146.6 (38.6) |
| TG (mg/dL) | 27547 | 120.0 [87.0, 168.0] | 4284 | 143.7 [105.0, 197.0] | 259 | 132.0 [96.0, 198.0] |
| FG (mg/dL) | 30281 | 94.0 [87.0, 103.0] | 3577 | 97.0 [89.0, 113.0] | 227 | 98.0 [90.0, 110.0] |
| FI (uU/mL) | 28751 | 8.0 [5.2, 12.5] | 3196 | 9.7 [6.2, 15.1] | 180 | 9.3 [6.1, 14.5] |
| Creatinine (mg/dL) | 20531 | 0.7 [0.7, 0.8] | 2307 | 0.7 [0.7, 0.8] | 124 | 0.8 [0.7, 0.9] |
| C-reactive protein (mg/dL) | 36210 | 2.6 [1.1, 5.5] | 4383 | 3.3 [1.5, 6.6] | 293 | 3.5 [1.7, 7.6] |
| Meta-GRS (SD) | 20374 | -0.1 (1.0) | 2314 | 0.2 (0.9) | 129 | 0.2 (0.9) |

N: total number of participants with non-missing covariate value. BMI: body mass index; SBP: systolic blood pressure; MET min/week: metabolic equivalent minutes per week of recreational activity; AHEI score: Alternative Healthy Eating Index score; CCB: calcium-channel blocker; RAS inhibitors: renin-angiotensin system inhibitors; LVHI: left ventricular hypertrophy index; LVHI\_CAT: Left Ventricular Hypertrophy Index > 120 (1 vs 0); NOVA5: Novacode 5 for Myocardial Infarction / Ischemia; MC\_MI: MI by Minnesota Code; HDLC: high-density lipoprotein cholesterol; LDLC: low-density lipoprotein cholesterol; TG: triglycerides; FG: fasting glucose; FI: fasting insulin; Education level was modeled as a continuous variable ranging from 1 to 11; Family income was modeled as a continuous variable ranging from 1 to 9; Meta-GRS: standard deviation of the normalized weighted genetic risk score of 1.7 million polymorphic susceptibility variants for CHD. Descriptive statistics for continuous variables that follow normal distribution, continuous variables that do not follow normal distribution, and binary variables were expressed as mean (standard deviation), median (first quartile, third quartile), and percentage respectively.

Our statistically most significant predictor of a fatal outcome at the time of an incident CHD event was age (**Table 2**). For each five-year increase in age, women were at a substantially increased risk for both fatal and non-fatal initial presentations of CHD. However, the increase in the risk for a fatal presentation was 46% greater (95% Confidence interval [CI] 35% to 58%) than that observed for a non-fatal presentation (**Table 2**).

**Table 2.**
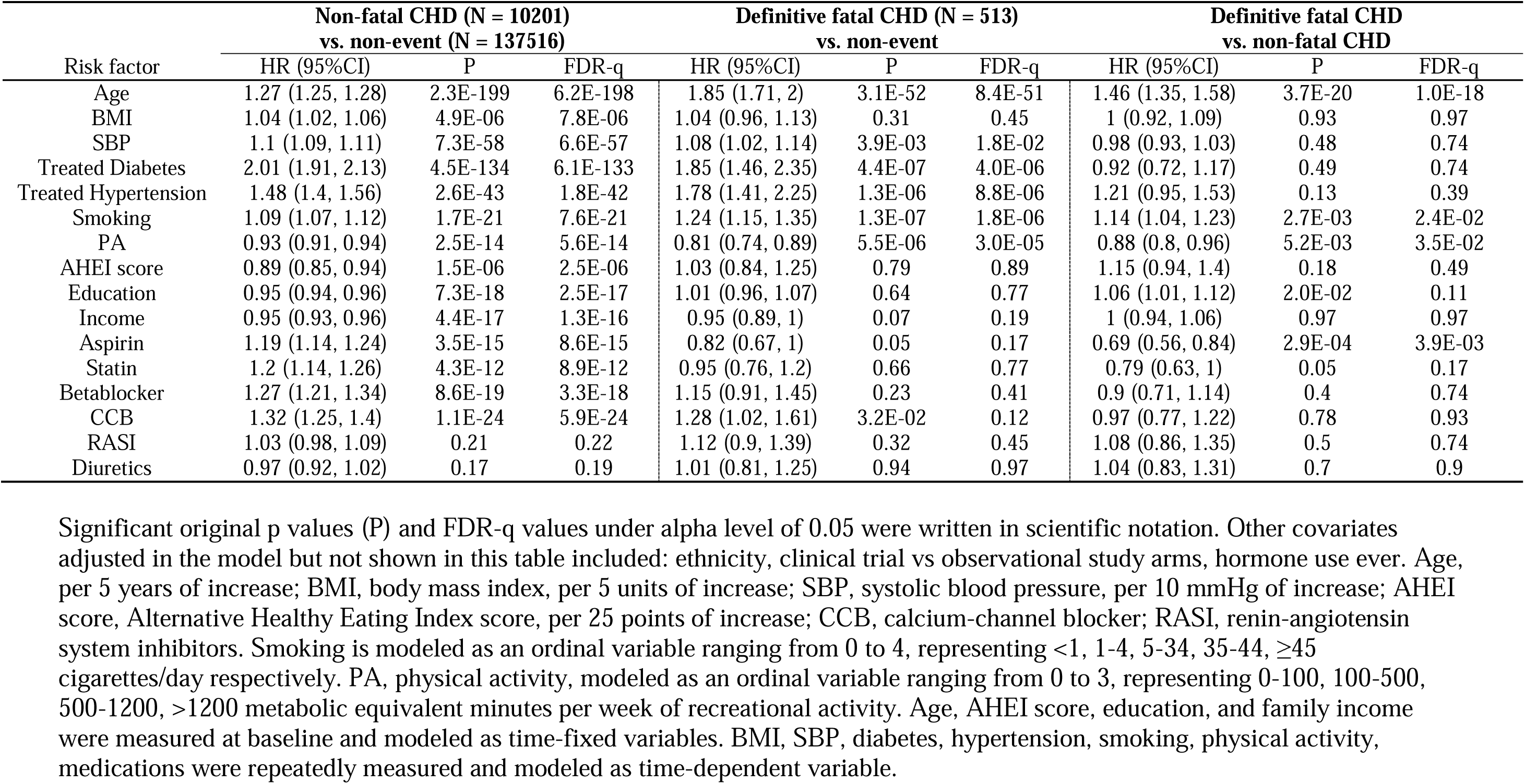
Association between premorbid predictors and each of the two types of incident CHD events derived from a joint multivariate Cox model, followed by the relative hazard ratio for a fatal vs. a non-fatal CHD event, of all participants after imputing missingness.

| Risk factor | Non-fatal CHD (N = 10201)<br>vs. non-event (N = 137516) |  |  | Definitive fatal CHD (N = 513)<br>vs. non-event |  |  | Definitive fatal CHD<br>vs. non-fatal CHD |  |  |
| --- | --- | --- | --- | --- | --- | --- | --- | --- | --- |
|  | HR (95%CI) | P | FDR-q | HR (95%CI) | P | FDR-q | HR (95%CI) | P | FDR-q |
| Age | 1.27 (1.25, 1.28) | 2.3E-199 | 6.2E-198 | 1.85 (1.71, 2) | 3.1E-52 | 8.4E-51 | 1.46 (1.35, 1.58) | 3.7E-20 | 1.0E-18 |
| BMI | 1.04 (1.02, 1.06) | 4.9E-06 | 7.8E-06 | 1.04 (0.96, 1.13) | 0.31 | 0.45 | 1 (0.92, 1.09) | 0.93 | 0.97 |
| SBP | 1.1 (1.09, 1.11) | 7.3E-58 | 6.6E-57 | 1.08 (1.02, 1.14) | 3.9E-03 | 1.8E-02 | 0.98 (0.93, 1.03) | 0.48 | 0.74 |
| Treated Diabetes | 2.01 (1.91, 2.13) | 4.5E-134 | 6.1E-133 | 1.85 (1.46, 2.35) | 4.4E-07 | 4.0E-06 | 0.92 (0.72, 1.17) | 0.49 | 0.74 |
| Treated Hypertension | 1.48 (1.4, 1.56) | 2.6E-43 | 1.8E-42 | 1.78 (1.41, 2.25) | 1.3E-06 | 8.8E-06 | 1.21 (0.95, 1.53) | 0.13 | 0.39 |
| Smoking | 1.09 (1.07, 1.12) | 1.7E-21 | 7.6E-21 | 1.24 (1.15, 1.35) | 1.3E-07 | 1.8E-06 | 1.14 (1.04, 1.23) | 2.7E-03 | 2.4E-02 |
| PA | 0.93 (0.91, 0.94) | 2.5E-14 | 5.6E-14 | 0.81 (0.74, 0.89) | 5.5E-06 | 3.0E-05 | 0.88 (0.8, 0.96) | 5.2E-03 | 3.5E-02 |
| AHEI score | 0.89 (0.85, 0.94) | 1.5E-06 | 2.5E-06 | 1.03 (0.84, 1.25) | 0.79 | 0.89 | 1.15 (0.94, 1.4) | 0.18 | 0.49 |
| Education | 0.95 (0.94, 0.96) | 7.3E-18 | 2.5E-17 | 1.01 (0.96, 1.07) | 0.64 | 0.77 | 1.06 (1.01, 1.12) | 2.0E-02 | 0.11 |
| Income | 0.95 (0.93, 0.96) | 4.4E-17 | 1.3E-16 | 0.95 (0.89, 1) | 0.07 | 0.19 | 1 (0.94, 1.06) | 0.97 | 0.97 |
| Aspirin | 1.19 (1.14, 1.24) | 3.5E-15 | 8.6E-15 | 0.82 (0.67, 1) | 0.05 | 0.17 | 0.69 (0.56, 0.84) | 2.9E-04 | 3.9E-03 |
| Statin | 1.2 (1.14, 1.26) | 4.3E-12 | 8.9E-12 | 0.95 (0.76, 1.2) | 0.66 | 0.77 | 0.79 (0.63, 1) | 0.05 | 0.17 |
| Betablocker | 1.27 (1.21, 1.34) | 8.6E-19 | 3.3E-18 | 1.15 (0.91, 1.45) | 0.23 | 0.41 | 0.9 (0.71, 1.14) | 0.4 | 0.74 |
| CCB | 1.32 (1.25, 1.4) | 1.1E-24 | 5.9E-24 | 1.28 (1.02, 1.61) | 3.2E-02 | 0.12 | 0.97 (0.77, 1.22) | 0.78 | 0.93 |
| RASI | 1.03 (0.98, 1.09) | 0.21 | 0.22 | 1.12 (0.9, 1.39) | 0.32 | 0.45 | 1.08 (0.86, 1.35) | 0.5 | 0.74 |
| Diuretics | 0.97 (0.92, 1.02) | 0.17 | 0.19 | 1.01 (0.81, 1.25) | 0.94 | 0.97 | 1.04 (0.83, 1.31) | 0.7 | 0.9 |

Significant original p values (P) and FDR-q values under alpha level of 0.05 were written in scientific notation. Other covariates adjusted in the model but not shown in this table included: ethnicity, clinical trial vs observational study arms, hormone use ever. Age, per 5 years of increase; BMI, body mass index, per 5 units of increase; SBP, systolic blood pressure, per 10 mmHg of increase; AHEI score, Alternative Healthy Eating Index score, per 25 points of increase; CCB, calcium-channel blocker; RASI, renin-angiotensin system inhibitors. Smoking is modeled as an ordinal variable ranging from 0 to 4, representing <1, 1-4, 5-34, 35-44, ≥45 cigarettes/day respectively. PA, physical activity, modeled as an ordinal variable ranging from 0 to 3, representing 0-100, 100-500, 500-1200, >1200 metabolic equivalent minutes per week of recreational activity. Age, AHEI score, education, and family income were measured at baseline and modeled as time-fixed variables. BMI, SBP, diabetes, hypertension, smoking, physical activity, medications were repeatedly measured and modeled as time-dependent variable.

Our second statistically most significant predictor of a fatal outcome at the time of incident CHD was aspirin use (**Table 2**). Women who took aspirin and developed a CHD event in WHI had a 31% (95% CI: 16 to 44%) lower risk of dying from their initial presentation of CHD than women who did not take aspirin (**Table 2**). Women taking aspirin, statins, beta-blockers, and CCBs identified were at higher increased risk of non-fatal CHD ranging from 19 to 32%, consistent with confounding by indication (**Table 2**). Similar hazard ratios for fatal CHD were observed with the use of beta-blockers and CCBs.

Our third statistically most significant predictor was smoking (**Table 2**, **Figure 2A,2C**). The relative hazard ratio of fatal vs. non-fatal CHD outcomes was similar for women reporting 5-14, 15-24, and 25-34 cigarettes per day. Thus, these exposure groups were combined. Women who smoked within a range of 5 to 34 cigarettes/day (weighted average of ∼16 cigs/day) suffered from an increase in the risk of both fatal and non-fatal initial presentations of CHD when compared to women who were non-smokers. However, the increase in risk for a fatal presentation was 28% greater (95% CI: 5% to 56%) than that observed for a non-fatal presentation (**Figure 2C**). Women with higher levels of smoking also observed a greater increase in the risk of fatal events compared to non-fatal events, with a ∼14% greater increase (95% CI: 4 to 23%) of risk observed per increasing level of smoking (**Table 2**).

**Figure 2.**
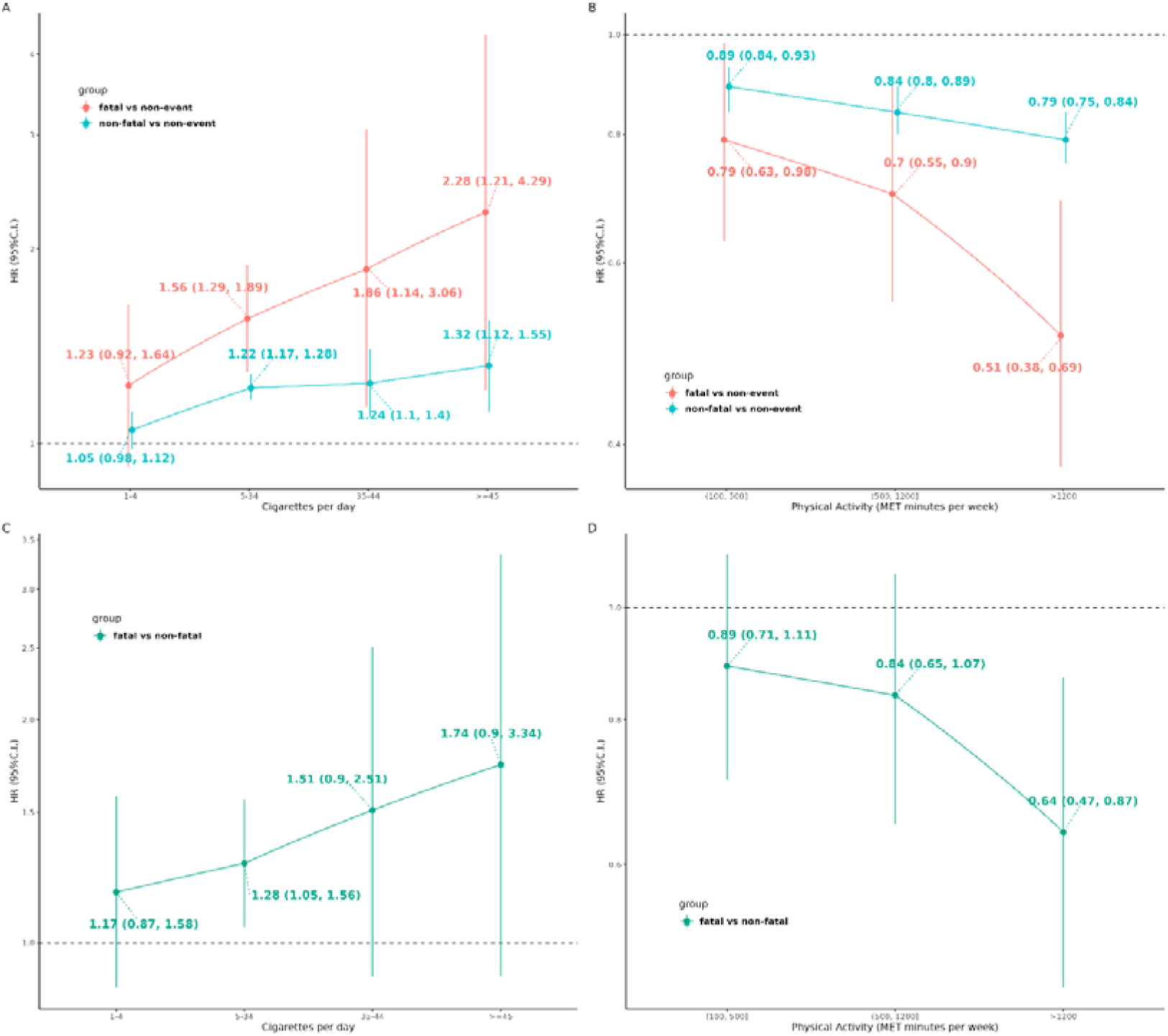
Hazard ratio of smoking (A and C) and physical activity (B and D) with each of the two subtypes of presentation of incident CHD, fatal (red curve), non-fatal (blue curve), and relative HR (green curve) using a joint multivariate Cox model. The level of physical activity was categorized into four groups: sedentary: ≤ 100 MET minutes/week (min/wk); low PA level: >100 and ≤500 MET min/wk; moderate PA: >500 and ≤1200 MET min/wk; high PA: >1200 MET min/wk. The level of smoking was categorized into five groups: <1, 1-4, 5-34, 35-44, ≥45 cigarettes/day)

Our fourth statistically most significant predictor was recreational physical activity (PA) (**Table 2**, **Figure 2B,2D**). Women reporting a high amount of PA benefited from a reduction in the risk of both fatal and non-fatal initial presentations of CHD when compared to sedentary women. However, the reduction in risk for a fatal presentation was 36% greater than that observed for a non-fatal presentation (**Figure 2D**). Women with intermediate levels of PA also observed a greater reduction in the risk of fatal events compared to non-fatal events, with a 16% greater reduction of risk observed per increasing level of PA (**Table 2**).

Next, we analyzed subgroups of women with ECG measures, blood biomarkers, and genetic data. Summary statistics of all the ECG measurements at baseline are provided in the supplement (**Table S3)**. Due to a high correlation, we excluded a subset of ECG measurements from our analyses (**Table S4**). Two predictors were significant at an FDR<0.05. Both were measures related to the baseline ECG. The most significant predictor was the Cornell voltage product, a measure that correlates with left ventricular mass on echocardiography (**Table S5**)^22^. The increase in risk for a fatal presentation was 17% (95% CI: 7 to 28%) greater than that observed for a non-fatal presentation per one SD increase in the Cornell Voltage product. Similar magnitude of increased risk was observed for the QRS duration (17%, 95% CI: 6 to 29%) (**Table S5**). We found no difference in the risk of dying from the initial presentation of CHD associated with a widely validated polygenic risk score (meta-GRS), LDL, HDL, triglycerides, creatinine, hs-CRP, and heart rate.

Lastly, we estimated cumulative absolute probabilities of presenting with an incident CHD event that was fatal over 20 years of follow in various subgroups as well as ratios of these probabilities between subgroups. For women aged 55, 65, and 75 years at baseline with a healthy lifestyle (no smoking and > 1200 MET min/week of PA), the cumulative probability was 0.1%, 0.37%, and 1.2%, respectively, while it increased to 0.4%, 1.3%, and 4.1% for women with an unhealthy lifestyle (smoking 35 to 44 cigarettes/day and < 100 MET min/week of PA) (**Figure 3a**, **Table S6**). The analogous cumulative probabilities for presenting with a non-fatal incident CHD event were 3.7%, 5.8%, and 9.1% for a healthy lifestyle and 6.0%, 9.4%, and 14.4% for an unhealthy lifestyle (**Figure 3a**, **Table S6**). Overall, the ratio of fatal to total (fatal and non-fatal) initial presentations of CHD for women with a healthy lifestyle compared to women with an unhealthy lifestyle decreased from 6.1% to 2.9%, 12.0% to 6.0%, and 22.3% to 11.9% for women aged 55, 65, 75 years, respectively (**Table S6**). Taking aspirin reduced the cumulative probability of fatal-CHD from 1.2% to 1.0% and 4.1% to 3.3% among 75-year women with a healthy lifestyle and an unhealthy lifestyle, respectively, and decreased the ratio of fatal to total initial presentations from 22.3% to 16.4% and 11.9% to 8.5% (**Figure 3b**, **Table S6**).

**Figure 3.**
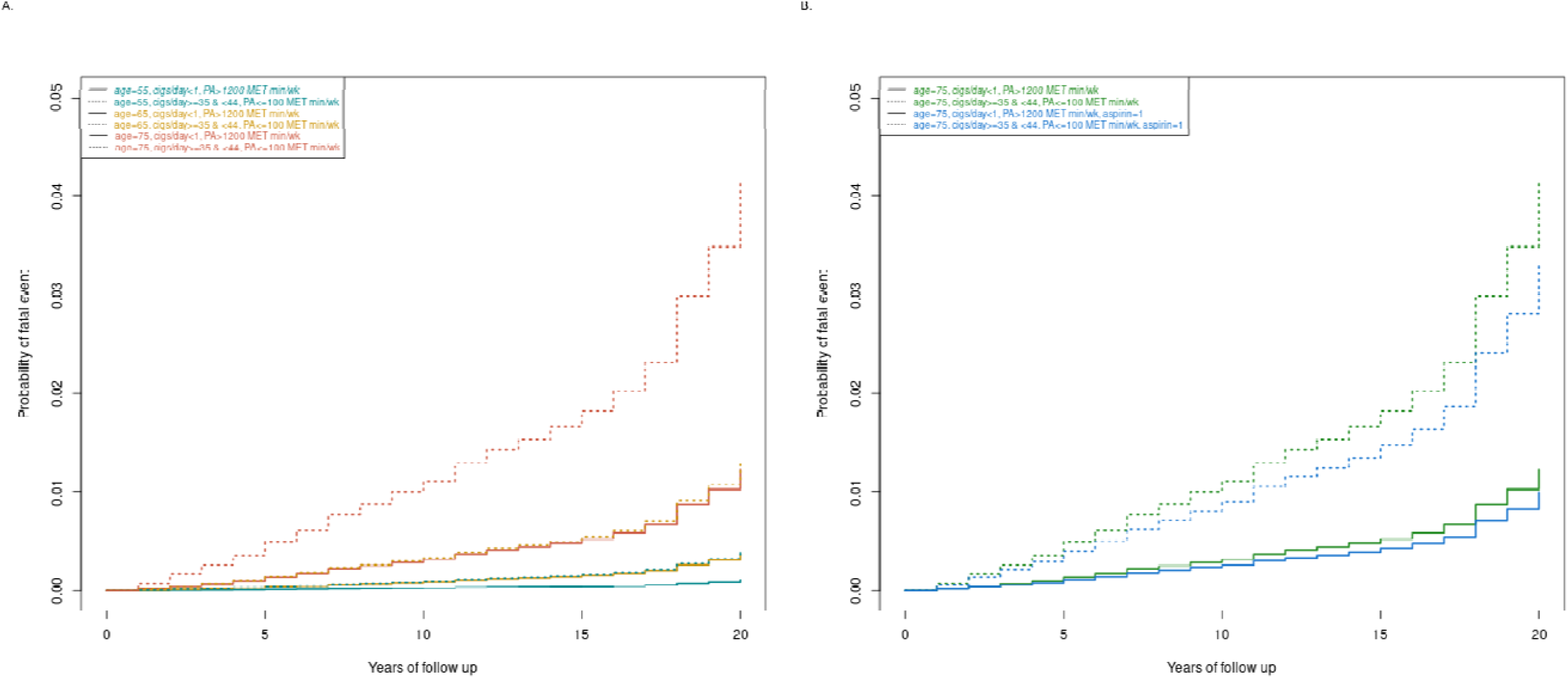
Estimated cumulative absolute risk of a fatal presentation of CAD over a 20 year follow up for subgroup of women using a joint multivariate Cox model. The cumulative probability of a fatal incident CHD event was modeled taking into consideration the competing risk of a non-fatal CHD event. Left graph compares the hazard function of women aged 55, 65, and 75 at baseline with and without healthy lifestyles as defined by no smoking and > 1200 MET min/week of PA vs. smoking 35 to 44 cigarettes/day and < 100 MET min/week of PA. Right graph compares women aged 75 at baseline with and without use of aspirin in addition to women with and without a healthy lifestyle.

## Discussion

We conducted to our knowledge the largest study to date of the premorbid predictors of the mode of presentation of CHD by leveraging data collected from the largest and longest-running cohort study of post-menopausal women in the United States. We summarize our findings thematically around four key observations.

First, we found older age, physical activity, smoking, aspirin use, and two ECG-derived measures (Cornell Voltage as a proxy for LV hypertrophy/mass and QRS duration) to be independent predictors of a fatal outcome among participants suffering their first-ever CHD event. While the findings for age and LVH are consistent with previously published findings, the remaining associations have not been previously reported in this specific context. The association between a prolonged QRS and a fatal outcome is plausible given a common cause of such prolongations is coronary artery disease^23,24^. Thus, these measures may be serving as markers of advanced subclinical CHD^23,24^. These prolongations may also be predisposing to fatal arrhythmic events in the setting of acute coronary syndromes^23,24^. Importantly, the non-ECG associations involve highly modifiable risk factors which have the potential to profoundly affect the case-fatality rate of incident CHD if they are optimally embraced at the population level.

Second, we could not replicate diabetes as an independent risk factor for an elevated short term (<28 days) case-fatality rate of CHD as was observed in the CHS study^8^ despite diabetes roughly doubling the risk of both incident and fatal CHD in WHI. To explain this discrepancy, we note that follow up and events for CHS accrued during the late 1980s and early 1990s while for WHI they accrued during the late 1990s, 2000s, and early 2010s. During this time, multiple other studies of cohorts in Finland, Germany, and Australia^25–27^ have shown substantial reductions in the case-fatality rates among subjects with and without diabetes including one study showing diabetes was no longer a predictor of increased case-fatality rate by 2010^27^. Differences in other demographic and clinical characteristics between CHS and WHI including baseline age distribution, prevalence and severity of diabetes, and education status^26^ may have also decreased power to detect this association. For example, the baseline prevalence of diabetes in CHS was 25.2%^8^ while it was only 3.6% in WHI.

Third, we found a healthy lifestyle at baseline (defined as no smoking and > 1200 MET min/week of PA) when compared to an unhealthy lifestyle (smoking 35 to 44 cigarettes/day and < 100 MET min/week of PA) to be associated with a reduction in the cumulative probability of dying from one’s initial presentation of CHD equivalent to the reduction observed among groups of women separated by approximately a decade in age. The effect of aspirin on lowering this risk was not as pronounced but still significant for all post-menopausal women.

Lastly, we note that probability of dying from initial presentation of CHD had a more rapid rate of increase relative to the probability of presenting with a non-fatal CHD over time, leading to an increasing absolute difference in the probability of dying from the initial presentation of CHD over time. This increasing difference became particularly evident in the second decade of follow-up. Thus, a healthy lifestyle at baseline is associated with a larger protection from a fatal incident CHD event late in life.

Strengths of our study include the long duration of follow-up, the availability of a very large number of carefully adjudicated incident fatal and non-fatal CHD events, and the repeated assessment of multiple exposures. Our baseline cohort produced over seven times the number of incident fatal CHD outcomes compared to the CHS study^8^. Our time-dependent analyses using a joint Cox model allowed us to optimally model the short-term effects of multiple exposures such as physical activity, smoking, and medications.

The limitations of this study should be considered carefully. First, our analysis of the most used classes of cardiovascular drugs was disadvantaged by the presence of confounding by indication given prior clinical trials have unequivocally established the protective effects of aspirin and statins, as well as antihypertensives, on the risk of incident CHD^28^. This type of confounding is common in observational studies reporting on the intended effects of pharmacological therapies and often cannot be corrected through covariate adjustment^29^. However, this confounding may be minimized or even eliminated when estimating the difference in risk reduction between fatal and non-fatal CHD events if it is in the same direction, and present to an equal degree, for both types of outcomes. Furthermore, the protective effect observed against fatal outcomes is biologically plausible given aspirin’s established role in limiting the extent of occlusive thrombosis that follows a plaque rupture. Second, we examined only post-menopausal women. We suspect that our findings do generalize to other populations although further study in large cohorts is needed to definitively prove this hypothesis. Third, we had reduced power to detect differences in the hazard ratios between fatal and non-fatal presentations of disease for our assessment of biomarkers and genetic risk. These risk factors need to be further examined in other large cohorts before definitive conclusions can be made. Fourth, no systematic assessment of LV function was available in WHI making it impossible to compare our results to the CHS in this respect^8^. However, routine assessment of LV function in asymptomatic individuals is not recommended for risk stratification. Lastly, we could not adjust for in hospital treatments that may have influenced case-fatality rates of CHD such as time from symptom onset of ACS to hospital presentation for thrombolytic therapy or primary coronary intervention, as such information was not collected by WHI^12^. Older age, diabetes, and smoking have been linked to prehospital delays involving ACSs and may be contributing to our observed results^30^. We note that smoking has been associated with a *shorter* prehospital delay perhaps due to the widely publicized link between smoking and the risk of CHD^30^. Thus, the increased risk of fatal presentation we observed for smoking may be an underestimate of the true effect.

Our analysis yields three insights within the framework of existing evidence and prevention protocols. First, the American Heart Association recommends either 150 minutes of moderate-intensity aerobic PA per week or 75 minutes of vigorous PA per week for the maintenance of optimal cardiovascular health. These levels of PA translate to ∼750 MET minutes per week. However, more extensive health benefits have been observed with additional PA^31^. Our findings suggest that higher levels of aerobic PA will have a more profound beneficial effect on a woman’s chances of dying from an incident CHD event. Second, we note that despite an established monotonic increase in the risk of CHD between individuals who smoke five and individuals who smoke 30 cigarettes per day^32^, the benefit we observed with respect to fatal outcomes over non-fatal outcomes in this range was relatively flat. Thus, our findings suggest that even smoking lightly disproportionally increases a woman’s risk of dying from her initial CHD event. Third, the use of aspirin in the primary prevention of CHD has been controversial with the most recent statement recommending the potential use for individuals aged 40-59 years of age with a >10% 10-year risk of atherosclerotic cardiovascular disease^33^. However, these recommendations account for the elevated risk of bleeding associated with aspirin use in determining its overall risk: benefit ratio^33^. We did not examine bleeding side effects in this study but still provide evidence that the risk of dying from one’s initial presentation of CHD appears reduced with the use of aspirin.

## Conclusions

In conclusion, our study found that older age, smoking, and two ECG-related measures including LVH and prolonged QRS increased the risk of dying from a first ever presentation of CHD, while physical activity and aspirin reduced this risk. This information may be used as an effective motivator for women to optimize their lifestyle and significantly reduce their overall risk of CHD but also their chances of dying from their initial event, should they ever have one. These benefits are likely to extend to other populations including pre-menopausal women and men, but further study is needed to prove this hypothesis more definitively.

## Author Contributions

Designed the study: T. L. A.

Conducted the analysis: M-L. C, J. L. K. R. I., C. T., S. J., E. L. I. S., L. C. D.

Wrote the paper: M-L. C., J. L., T. L. A.

Funding and/or resources: M. L. S., M. D., X. X.

Provided final review: All authors.

## Acknowledgements

We acknowledge the contributions of the following past and present key WHI investigators:

**Program Office:** (National Heart, Lung, and Blood Institute, Bethesda, Maryland) Jacques Rossouw, Shari Ludlam, Joan McGowan, Leslie Ford, and Nancy Geller

**Clinical Coordinating Center**: (Fred Hutchinson Cancer Research Center, Seattle, WA) Garnet Anderson, Ross Prentice, Andrea LaCroix, and Charles Kooperberg

**Investigators and Academic Centers**: (Brigham and Women’s Hospital, Harvard Medical School, Boston, MA) JoAnn E. Manson; (MedStar Health Research Institute/Howard University, Washington, DC) Barbara V. Howard; (Stanford Prevention Research Center, Stanford, CA) Marcia L. Stefanick; (The Ohio State University, Columbus, OH) Rebecca Jackson; (University of Arizona, Tucson/Phoenix, AZ) Cynthia A. Thomson; (University at Buffalo, Buffalo, NY) Jean Wactawski-Wende; (University of Florida, Gainesville/Jacksonville, FL) Marian Limacher; (University of Iowa, Iowa City/Davenport, IA) Jennifer Robinson; (University of Pittsburgh, Pittsburgh, PA) Lewis Kuller; (Wake Forest University School of Medicine, Winston-Salem, NC) Sally Shumaker; (University of Nevada, Reno, NV) Robert Brunner

**Women’s Health Initiative Memory Study**: (Wake Forest University School of Medicine, Winston-Salem, NC) Mark Espeland

## Data availability Statement

Qualified investigators may gain access to the WHI data through the WHI data coordinating center after securing approval of a paper proposal as described here https://www.whi.org/propose-a-paper. Data is also accessible through BiolinC at https://biolincc.nhlbi.nih.gov/studies/whi_ctos/ and through dbGAP at https://www.ncbi.nlm.nih.gov/projects/gap/cgi-bin/study.cgi?study_id=phs000200.v12.p3 The 1000G reference panel used for joint imputation can be accessed through https://www.ncbi.nlm.nih.gov/projects/gap/cgi-bin/study.cgi?study_id=phs000746.v1.p3.

