## Supplemental Text and Tables for "Premorbid predictors of death at initial presentation of coronary heart disease in the Women’s Health Initiative study"

Supplemental Material

### **Supplemental Methods**

#### Schedule of Assessment

The complete schedule of assessment for all characteristics were described on the WHI website (<https://www.whi.org/researchers/data/Documents/Participant%20Characteristics%20over%20Time.pdf>). The schedule of assessment for time-dependent variables in our study was summarized in **Supplementary Table 1**.

#### Exposure Assessment

At the in-person baseline visit, trained personnel also measured participant’s height, weight, blood pressure, and drew blood that was banked for future biomarker studies. We used the following medications to determine whether participants were exposed to aspirin, statins, diuretics, beta-blockers, calcium-channel blockers, and inhibitors of the renin-angiotensin system (RAS).

Smoking status and amount of smoking was coded as an ordinal variable ranging from 0 to 4, representing a self-report of smoking <1, 1-4, 5-34, 35-44, ≥ 45 cigarettes/day, respectively. The WHI physical activity (PA) questionnaire documented the frequency and duration of walking, and other mild, moderate, and strenuous types of physical activities. Using a standardized classification of the energy expenditure associated with various types of PAs^1^, we calculated a weekly energy expenditure for each woman of total PA in metabolic equivalents (MET) minutes per week. This score has been previously shown to have a high test-retest reliability, to be a valid measure of PA based on two referent methods of assessment, and to be inversely associated with the risk of incident coronary events^2^. We further categorized the levels of PA into four groups: sedentary: ≤100 MET · min/wk; low PA level: >100 and ≤500 MET · min/wk; moderate PA: >500 and ≤1200 MET · min/wk; high PA: >1200 MET · min/wk^3^.

We estimated the quality of participant’s diet at baseline by calculating the Alternative Healthy Eating Index (AHEI) 2010 score from the food frequency questionnaire.^4^ The score includes 11 dietary components adjusted for energy using the residuals method: 1.) vegetables 2.) fruits 3.) whole grains 4.) sugar-sweetened beverages & fruit juice 5.) nuts and legumes 6.) red & processed meat 7.) trans-fat 8.) long-chain (n-3) fats (eicosapentaenoic + docosahexaenoic acids) 9.) n-3 and n-6 polyunsaturated fatty acids 10.) sodium 11.) alcohol. A maximum of 110 points could be achieved, with higher scores indicating higher diet quality.

Education level was modeled as a continuous variable ranging from 1 to 11 (1: Didn’t go to school; 2: Grade school 1-4 years; 3: Grade school 5-8 years; 4: Some high school 9-11 years; 5: High school diploma or GED; 6: Vocational or training school; 7: Some college or Associate degree; 8: College graduate or Baccalaureate Degree; 9: Some post-graduate or professional;10: Master’s Degree 11: Doctoral Degree (Ph.D, M.D., J.D., etc.). Family income was modeled as a continuous variable ranging from 1 to 8 (1: Less than $10,000; 2: $10,000 to $19,999; 3: $20,000 to $34,999; 4: $35,000 to $49,999; 5: $50,000 to $74,999; 6: $75,000 to $99,999; 7: $100,000 to $149,999; 8: $150,000 or more).

Standard 12-lead electrocardiograms were obtained at baseline for all trial participants. ECGs were coded by the Epidemiological Cardiology Research Center using Minnesota and Novacode coding.^5,6^ The MAC PC electrocardiograph (GE Healthcare, Waukesha, Wisconsin) was used to record the electrocardiograms and transmit them to the electrocardiography center at Wake Forest University in North Carolina for diagnostic classification. All electrocardiograms were given a quality grade to ensure adequate performance by the technicians. Quality assurance was enhanced by implementation of computerized and semiautomatic procedures.

A subset of close to 30,000 women including approximately 19,000 women of European descent underwent genome wide genotyping through one of six ancillary studies (Hip Fracture GWAS-BA23, SHARe-M5, GARNET-M13, WHIMS+-W63, GECCO-AS224, MOPMAP-AS264). The studies used a variety of high-density genotyping arrays from either Illumina or Affymetrix. The data were formally harmonized through imputation to the 1000 genomes reference panel of SNPs. From this imputed data, we extracted dosages for ~1.7 million single nucleotide polymorphisms (SNPs) included in a widely validated polygenic risk score (PGS) for clinical coronary artery disease (metaGRS) and calculated the score for an individual in the typical approach as a weighted sum of the number of high risk alleles. We modeled the allele associated with increased risk of CAD as the effect allele, and the weighted PGS was calculated by multiplying the allele score (0 to 2) for each SNP in WHI by the SNPs’s beta coefficient and summing the products across all the SNPs. The PGS was standardized so that it followed the normal distribution with mean 0 and variance 1.

#### Statistical Analysis

In a joint Cox model, the hazard functions for event *k* (*k*=1, 2) are denoted as λ*_k_* (t), and the hazard ratio (HR) associated with a unit increase in risk factors, X, for the development of the event 1 and event 2 are denoted as exp (β_1_) and exp (β_2_). If we let β_1_ ${=\beta}^{*}$, and β_2_ ${=\beta}^{*}+\theta$, according to the cox model, the hazard function can be reparameterized as:

λ*_k_* (t)= λ_0_*_k_* (t) exp ($\beta^{*}X+\theta I_{k=2}X$)

where $I_{k=2}$ equals to 1 if the event time is for event 2 and equals to 0 otherwise. Thus, an estimate of θ provides an estimate of β_1_ - β_2_ and a test of β_1=_ β_2_ is equivalent to a test of θ=0, which can be conducted with a Wald test. Therefore, using joint Cox model, HRs for the association of risk factors with two subtypes of CHD cases were estimated simultaneously and the differences in HRs were assessed. The joint Cox model is less susceptible to bias in comparing exposure-disease associations across disease subtypes in prospective cohort studies than polytomous logistic regression when the baseline hazards for the different presentations of disease are non-proportional^7^. The status was modeled as 1 for the two subtypes of cases and modeled as 0 for non-cases. The time of follow up for the two subtypes of cases was from enrollment to the first occurrence of a CHD event. The time of follow up for non-cases who were not in MRC, and non-cases who were enrolled in MRC were from enrollment to the end of Extension 1, and from enrollment to the end of Extension 2 respectively.

We performed sensitivity analysis using the main model only among individuals with complete, non-missing measurements of all the covariates included in the main analyses (N = 137959).

We assessed for the presence of confounding by indication for all major classes of cardiovascular disease drugs by running marginal structural models to assess the effect of this modelling on the associations with medication.^8^

### **Supplemental Results**

Sensitivity analysis limited to the subgroup of 137, 959 women with complete measurements of covariates included in our main analysis were consistent with our analyses of the full cohort after imputation of missing values. (**Supplemental Table 2**).

Marginal structural modelling did not materially change the observed risks (details not shown).

**Table S1**. Participant characteristics collected during follow-up

| variables | cohort | clinical trials and observational studies collection year (1993-2005) | | | | | | | | | | | extension study collection year (2005-2010) | | | | | extension study collection year (2010-2015) | | | | |
| --- | --- | --- | --- | --- | --- | --- | --- | --- | --- | --- | --- | --- | --- | --- | --- | --- | --- | --- | --- | --- | --- | --- |
|  |  | B | 1 | 2 | 3 | 4 | 5 | 6 | 7 | 8 | 9 | C | 1 | 2 | 3 | 4 | 5 | 1 | 2 | 3 | 4 | 5 |
| BMI | CT | Y | Y | Y | Y | Y | Y | Y | Y | Y | Y |  |  |  |  |  |  |  |  |  |  |  |
|  | OS | Y |  |  | Y |  |  |  |  |  |  |  |  |  |  |  |  |  |  |  |  |  |
| SBP | CT | Y | Y | Y | Y | Y | Y | Y | Y | Y | Y |  |  |  |  |  |  |  |  |  |  |  |
|  | OS | Y |  |  | Y |  |  |  |  |  |  |  |  |  |  |  |  |  |  |  |  |  |
| diabetes | CT | Y | Y | Y | Y | Y | Y | Y | Y | Y | Y |  | Y | Y | Y | Y | Y | Y | Y | Y | Y | Y |
|  | OS | Y | Y | Y | Y | Y | Y | Y | Y | Y | Y |  | Y | Y | Y | Y | Y | Y | Y | Y | Y | Y |
| hypertension | CT | Y | Y | Y | Y | Y | Y | Y | Y | Y | Y | Y | Y | Y | Y | Y | Y | Y | Y | Y | Y | Y |
|  | OS | Y | Y | Y | Y | Y | Y | Y | Y | Y | Y |  | Y | Y | Y | Y | Y | Y | Y | Y | Y | Y |
| smoking | CT | Y | Y |  | Y |  |  | Y |  |  | Y |  |  |  |  |  |  |  | Y |  |  |  |
|  | OS | Y | Y |  | Y | Y | Y | Y | Y | Y |  |  |  |  |  |  |  |  | Y |  |  |  |
| PA | CT | Y | Y |  | Y |  |  | Y |  |  | Y |  |  |  |  |  |  |  | Y |  |  |  |
|  | OS | Y |  |  | Y | Y | Y | Y | Y | Y |  |  |  |  |  |  |  |  | Y |  |  |  |
| medications | CT | Y | Y |  | Y |  |  | Y |  |  | Y |  |  |  |  | Y |  |  |  |  |  |  |
|  | OS | Y |  |  | Y |  |  |  |  |  |  |  |  |  |  | Y |  |  |  |  |  |  |

Note: B, baseline; C, closeout; CT, clinical trial; OS, observational study; BMI, body mass index; SBP, systolic blood pressure; PA, physical activity

**Table S2. Sensitivity Analysis. Association between premorbid predictors and each of the two types of incident CHD events derived from a joint multivariate Cox model, followed by the relative hazard ratio for a fatal vs. a non-fatal CHD event, using non-imputed, complete case data.**

|  | **Non-fatal CHD (N = 9534)**  **vs. non-event (N = 127955)** | | | **Definitive fatal CHD (N = 470)**  **vs. non-event** | | | **Definitive fatal CHD**  **vs. non-fatal CHD** | | |
| --- | --- | --- | --- | --- | --- | --- | --- | --- | --- |
| Risk factor | HR (95%CI) | P | FDR-q | HR (95%CI) | P | FDR-q | HR (95%CI) | P | FDR-q |
| Age | 1.27 (1.25, 1.29) | 2.6E-186 | 7.0E-185 | 1.82 (1.67, 1.97) | 2.1E-46 | 5.7E-45 | 1.44 (1.32, 1.56) | 2.1E-17 | 5.7E-16 |
| BMI | 1.04 (1.02, 1.06) | 6.4E-06 | 1.0E-05 | 1.05 (0.96, 1.15) | 0.25 | 0.42 | 1.01 (0.92, 1.11) | 0.81 | 0.91 |
| SBP | 1.1 (1.09, 1.11) | 4.9E-55 | 4.4E-54 | 1.08 (1.02, 1.14) | 5.4E-03 | 2.4E-02 | 0.98 (0.93, 1.04) | 0.52 | 0.77 |
| Treated Diabetes | 2.01 (1.9, 2.13) | 6.9E-125 | 9.3E-124 | 1.88 (1.47, 2.4) | 4.5E-07 | 4.0E-06 | 0.94 (0.73, 1.2) | 0.6 | 0.77 |
| Treated Hypertension | 1.47 (1.39, 1.56) | 2.3E-39 | 1.6E-38 | 1.85 (1.44, 2.36) | 1.0E-06 | 6.7E-06 | 1.26 (0.98, 1.62) | 0.08 | 0.27 |
| Smoking | 1.1 (1.08, 1.12) | 5.6E-22 | 2.5E-21 | 1.25 (1.15, 1.36) | 1.5E-07 | 2.0E-06 | 1.14 (1.05, 1.24) | 3.0E-03 | 2.4E-02 |
| PA | 0.93 (0.91, 0.94) | 1.3E-13 | 2.9E-13 | 0.8 (0.73, 0.88) | 3.9E-06 | 2.1E-05 | 0.87 (0.79, 0.96) | 3.6E-03 | 2.4E-02 |
| AHEI score | 0.89 (0.85, 0.93) | 7.3E-07 | 1.3E-06 | 1.11 (0.91, 1.36) | 0.3 | 0.42 | 1.25 (1.02, 1.54) | 3.3E-02 | 0.13 |
| Education | 0.95 (0.94, 0.96) | 2.8E-16 | 8.4E-16 | 0.99 (0.94, 1.05) | 0.8 | 0.89 | 1.04 (0.99, 1.1) | 0.14 | 0.32 |
| Income | 0.94 (0.93, 0.96) | 1.1E-16 | 3.4E-16 | 0.95 (0.89, 1.01) | 0.09 | 0.27 | 1 (0.94, 1.07) | 0.98 | 0.98 |
| Aspirin | 1.19 (1.14, 1.24) | 2.6E-14 | 7.0E-14 | 0.85 (0.7, 1.05) | 0.13 | 0.29 | 0.72 (0.58, 0.89) | 1.9E-03 | 2.4E-02 |
| Statin | 1.21 (1.15, 1.28) | 1.9E-12 | 3.9E-12 | 1.01 (0.79, 1.27) | 0.96 | 0.96 | 0.83 (0.65, 1.06) | 0.14 | 0.32 |
| Betablocker | 1.28 (1.22, 1.36) | 7.4E-19 | 2.9E-18 | 1.13 (0.89, 1.43) | 0.33 | 0.42 | 0.88 (0.69, 1.12) | 0.29 | 0.6 |
| CCB | 1.33 (1.26, 1.41) | 6.2E-24 | 3.3E-23 | 1.26 (1, 1.59) | 0.05 | 0.19 | 0.95 (0.74, 1.21) | 0.66 | 0.81 |
| RASI | 1.05 (0.99, 1.1) | 0.11 | 0.13 | 1.12 (0.89, 1.41) | 0.32 | 0.42 | 1.07 (0.85, 1.35) | 0.56 | 0.77 |
| Diuretics | 0.96 (0.91, 1.02) | 0.17 | 0.18 | 1.03 (0.82, 1.29) | 0.82 | 0.89 | 1.07 (0.84, 1.35) | 0.59 | 0.77 |

Dataset limited to individuals who have complete measurement of covariates displayed above. Other covariates adjusted in the model but not shown in this table included: ethnicity, clinical trial vs observational study arms, hormone use ever. Age, per 5 years of increase; BMI, body mass index, per 5 units of increase; SBP, systolic blood pressure, per 10 mmHg of increase; AHEI score, Alternative Healthy Eating Index score, per 25 points of increase; CCB, calcium-channel blocker; RASI, renin-angiotensin system inhibitors. Smoking is modeled as an ordinal variable ranging from 0 to 4, representing <1, 1-4, 5-34, 35-44, ≥ 45 cigarettes/day respectively. PA, physical activity, modeled as an ordinal variable ranging from 0 to 3, representing 0-100, 100-500, 500-1200, >1200 metabolic equivalent minutes per week of recreational activity. Age, AHEI score, education, and family income were measured at baseline and modeled as time-fixed variables. BMI, SBP, diabetes, hypertension, smoking, physical activity, medications were repeatedly measured and modeled as time-dependent variables.

**Table S3. Summary of EKG measurement at baseline.**

|  | **Women who did not develop CHD  (N = 137516)** | | | **Women who developed non-fatal CHD  (N = 10201)** | | | **Women who developed definitive fatal CHD (N = 513)** | | |
| --- | --- | --- | --- | --- | --- | --- | --- | --- | --- |
| ECG Measures | N | Mean (SD) | Median (IQR) or Frequency (%) | N | Mean (SD) | Median (IQR) or Frequency (%) | N | Mean (SD) | Median (IQR) or Frequency (%) |
| BRADYCAR | 53499 |  | 2.6 | 4615 |  | 2.9 | 216 |  | 2.3 |
| MC_MI | 51667 |  | 2 | 4378 |  | 3.7 | 197 |  | 5.6 |
| VE_N | 53499 |  | 1.6 | 4615 |  | 2.1 | 216 |  | 2.3 |
| LVH_MINN | 52245 |  | 5.4 | 4429 |  | 8.6 | 198 |  | 11.1 |
| PROLO_PR | 53499 |  | 1.6 | 4615 |  | 2.4 | 216 |  | 2.8 |
| ECGBASENOVA | 46719 |  | 26.3 | 4010 |  | 37.8 | 146 |  | 41.1 |
| LVHI | 53132 | 81.0 (7.3) | 80.0 [76.0, 84.0] | 4584 | 83.4 (8.8) | 82.0 [78.0, 87.0] | 212 | 85.4 (13.1) | 82.0 [78.0, 88.0] |
| LVHI_CAT | 53132 |  | 0.2 | 4584 |  | 0.7 | 212 |  | 3.3 |
| NOVA5 | 51735 |  | 28.6 | 4383 |  | 40.2 | 197 |  | 44.7 |
| CV | 53499 | 1198.9 (485.7) | 1161.0 [863.0, 1483.0] | 4615 | 1317.9 (519.8) | 1263.0 [966.0, 1616.0] | 216 | 1416.1 (657.2) | 1317.0 [1006.2, 1695.0] |
| CV_QRS | 53499 | 106.1 (52.9) | 98.8 [72.2, 130.2] | 4615 | 118.7 (60.3) | 109.5 [81.0, 143.3] | 216 | 135.8 (92.5) | 115.9 [86.3, 150.9] |
| LVMI_NORM | 50644 | 141.1 (20.8) | 139.6 [126.2, 154.4] | 4215 | 145.6 (20.4) | 144.8 [131.5, 158.7] | 186 | 144.6 (19.3) | 144.3 [131.0, 155.5] |
| LVM_NORM | 50470 | 53.8 (8.8) | 53.2 [47.8, 59.1] | 4209 | 56.3 (9.1) | 55.8 [50.4, 61.6] | 186 | 56.0 (7.3) | 55.7 [51.0, 61.2] |
| PR_DUR | 53499 | 160.0 (25.5) | 158.0 [144.0, 174.0] | 4615 | 162.4 (28.3) | 160.0 [146.0, 178.0] | 216 | 164.2 (24.7) | 162.0 [146.0, 182.0] |
| QTC_DUR | 53499 | 418.8 (19.1) | 415.0 [407.0, 427.0] | 4615 | 422.0 (20.8) | 417.0 [408.0, 431.0] | 216 | 426.6 (31.6) | 417.5 [408.0, 436.0] |
| QT_IND | 53499 | 101.1 (4.5) | 100.0 [98.0, 103.0] | 4615 | 101.8 (4.9) | 101.0 [98.0, 104.0] | 216 | 102.7 (7.5) | 101.0 [98.8, 106.0] |
| QT_DUR | 53499 | 402.1 (30.7) | 400.0 [382.0, 420.0] | 4615 | 403.1 (33.0) | 402.0 [382.0, 424.0] | 216 | 403.4 (38.2) | 398.0 [378.0, 422.0] |
| QRS_DUR | 53499 | 87.1 (11.6) | 86.0 [80.0, 92.0] | 4615 | 88.4 (13.7) | 86.0 [80.0, 92.0] | 216 | 91.8 (18.6) | 87.0 [82.0, 94.0] |

BRADYCAR, Bradycardia, 0 vs 1; MC_MI, MI by Minnesota Code, 0 vs 1; VE_N, Ventricular etopic beats, 0 vs 1; LVH_MINN, LVH by Minnesota Code, 0 vs 1; PROLO_PR, Prolonged PR interval, 0 vs 1; ECGBASENOVA, ECG MI novacode, 0 vs 1; LVHI: left ventricular hypertrophy index; LVHI_CAT: Left Ventricular Hypertrophy Index > 120 (1 vs 0); NOVA5: Novacode 5 for Myocardial Infarction / Ischemia, no Q_ST abnormalities (0) vs any abnormality (1); CV, Cornell Voltage (uV); CV_QRS, Cornell Voltage Production (uV)= (Cornell Voltage x QRS Duration / 1000); LVMI_Norm, LeftVentricular Mass Index (%); LVM_Norm, Left Ventricular Mass (g); PR_DUR, PR Interval (ms); QTC_DUR, Bazett's Heart Rate-Corrected QT Interval (ms); QT_IND, Rautaharju's QT Prolongation Index (%); QT_DUR, QT Interval Duration (ms); QRS_DUR, QRS Interval Duration (ms).;

**Table S4. Correlation matrix of EKG measurement at baseline.**

|  | BRADYCAR | LVH MINN | LVMI NORM | LVM NORM | NOVA5 | QT_IND | CV | CV_QRS | PR _DUR | PROLO _PR | QRS _DUR | QTC _DUR | QT _DUR | VE_N | ECGBASENOVA |
| --- | --- | --- | --- | --- | --- | --- | --- | --- | --- | --- | --- | --- | --- | --- | --- |
| BRADYCAR | 1.00 |  |  |  |  |  |  |  |  |  |  |  |  |  |  |
| LVH_MINN | 0.01 | 1.00 |  |  |  |  |  |  |  |  |  |  |  |  |  |
| LVMI_NORM | -0.02 | 0.21 | 1.00 |  |  |  |  |  |  |  |  |  |  |  |  |
| LVM_NORM | -0.02 | 0.20 | 0.82 | 1.00 |  |  |  |  |  |  |  |  |  |  |  |
| NOVA5 | 0.00 | 0.10 | 0.19 | 0.19 | 1.00 |  |  |  |  |  |  |  |  |  |  |
| QT_IND | 0.15 | 0.06 | 0.16 | 0.13 | 0.12 | 1.00 |  |  |  |  |  |  |  |  |  |
| CV | 0.01 | 0.30 | 0.68 | 0.67 | 0.21 | 0.16 | 1.00 |  |  |  |  |  |  |  |  |
| CV_QRS | 0.02 | 0.30 | 0.67 | 0.64 | 0.20 | 0.19 | 0.97 | 1.00 |  |  |  |  |  |  |  |
| PR_DUR | 0.05 | 0.06 | 0.14 | 0.09 | 0.03 | 0.05 | 0.10 | 0.11 | 1.00 |  |  |  |  |  |  |
| PROLO_PR | 0.03 | 0.01 | 0.03 | 0.02 | 0.03 | 0.02 | 0.03 | 0.04 | 0.39 | 1.00 |  |  |  |  |  |
| QRS_DUR | 0.05 | 0.08 | 0.26 | 0.16 | 0.04 | 0.19 | 0.27 | 0.48 | 0.09 | 0.03 | 1.00 |  |  |  |  |
| QTC_DUR | -0.04 | 0.06 | 0.19 | 0.16 | 0.14 | 0.92 | 0.17 | 0.18 | 0.01 | 0.00 | 0.14 | 1.00 |  |  |  |
| QT_DUR | 0.38 | 0.03 | 0.00 | -0.02 | 0.03 | 0.62 | 0.05 | 0.09 | 0.12 | 0.05 | 0.19 | 0.29 | 1.00 |  |  |
| VE_N | -0.01 | 0.00 | 0.02 | 0.02 | 0.04 | 0.08 | 0.02 | 0.02 | 0.00 | 0.01 | 0.02 | 0.09 | 0.00 | 1.00 |  |
| ECGBASENOVA | 0.00 | 0.09 | 0.18 | 0.18 | 0.92 | 0.12 | 0.20 | 0.20 | 0.03 | 0.03 | 0.05 | 0.14 | 0.02 | 0.03 | 1.00 |

BRADYCAR, Bradycardia, 0 vs 1; LVH_MINN, LVH by Minnesota Code, 0 vs 1; LVMI_Norm, Left Ventricular Mass Index (%); LVM_Norm, Left Ventricular Mass (g); NOVA5: Novacode 5 for Myocardial Infarction / Ischemia, no Q_ST abnormalities (0) vs any abnormality (1); QT_IND, Rautaharju's QT Prolongation Index (%); CV, Cornell Voltage (uV); CV_QRS, Cornell Voltage Production (uV)= (Cornell Voltage x QRS Duration / 1000); PR_DUR, PR Interval (ms); PROLO_PR, Prolonged PR interval, 0 vs 1; QRS_DUR, QRS Interval Duration (ms); QTC_DUR, Bazett's Heart Rate-Corrected QT Interval (ms); QT_DUR, QT Interval Duration (ms); VE_N, Ventricular etopic beats, 0 vs 1; ECGBASENOVA, ECG MI novacode, 0 vs 1.

**Table S5.** **Subgroup Analysis. Association of ECG parameters, blood biomarker levels, and a genetic risk score with each of the two types of incident CHD events derived from a joint multivariate Cox model, followed by the relative hazard ratio for a fatal vs. a non-fatal CHD event, using imputed data.**

|  | **Non-fatal CHD vs non-event** | | | | **Definitive fatal CHD vs non-event** | | | | **Definitive fatal CHD vs non-fatal CHD** | | |
| --- | --- | --- | --- | --- | --- | --- | --- | --- | --- | --- | --- |
| Risk factor | N | HR (95%CI) | P | FDR-q | N | HR (95%CI) | P | FDR-q | HR (95%CI) | P | FDR-q |
| Bradycardia | 4615 | 1.11 (0.93, 1.32) | 0.24 | 0.24 | 216 | 0.86 (0.35, 2.09) | 0.73 | 0.73 | 0.77 (0.31, 1.91) | 0.57 | 0.82 |
| VE_N | 4615 | 1.17 (0.96, 1.44) | 0.12 | 0.13 | 216 | 1.21 (0.5, 2.96) | 0.67 | 0.73 | 1.03 (0.41, 2.58) | 0.94 | 1 |
| CV | 4615 | 1.08 (1.05, 1.11) | 2.7E-07 | 5.6E-07 | 216 | 1.26 (1.11, 1.44) | 4.9E-04 | 2.3E-03 | 1.17 (1.02, 1.33) | 2.4E-02 | 0.13 |
| CV_QRS | 4615 | 1.08 (1.05, 1.11) | 6.1E-08 | 1.4E-07 | 216 | 1.26 (1.16, 1.38) | 8.2E-08 | 1.9E-06 | 1.17 (1.07, 1.28) | 4.0E-04 | 9.2E-03 |
| LVH_MINN | 4429 | 1.23 (1.1, 1.37) | 1.8E-04 | 2.8E-04 | 198 | 1.64 (1.05, 2.55) | 3.0E-02 | 0.05 | 1.33 (0.84, 2.11) | 0.22 | 0.72 |
| LVMI_Norm | 4215 | 1.06 (1.02,1.11) | 5.2E-03 | 7.0E-03 | 186 | 1.16 (0.97, 1.4) | 0.11 | 0.15 | 1.09 (0.91, 1.32) | 0.35 | 0.73 |
| LVM_Norm | 4209 | 1.1 (1.07, 1.13) | 6.1E-10 | 1.6E-09 | 186 | 1.14 (1.05,1.24) | 1.4E-03 | 4.0E-03 | 1.04 (0.96, 1.13) | 0.33 | 0.73 |
| PR_DUR | 4615 | 1.02 (0.99,1.05) | 0.17 | 0.18 | 216 | 1.06 (0.95, 1.2) | 0.29 | 0.35 | 1.04 (0.92, 1.18) | 0.5 | 0.81 |
| PROLO_PR | 4615 | 1.24 (1.03, 1.5) | 2.6E-02 | 3.3E-02 | 216 | 1.25 (0.55, 2.84) | 0.6 | 0.69 | 1.01 (0.43, 2.34) | 0.99 | 1 |
| QRS_DUR | 4615 | 1.04 (1.02,1.07) | 1.8E-03 | 2.6E-03 | 216 | 1.22 (1.11,1.34) | 3.1E-05 | 3.6E-04 | 1.17 (1.06, 1.29) | 1.5E-03 | 1.7E-02 |
| QT_DUR | 4615 | 1.03 (1, 1.06) | 0.05 | 0.06 | 216 | 1.03 (0.88, 1.2) | 0.71 | 0.73 | 1 (0.86, 1.17) | 1 | 1 |
| QTC_DUR | 4615 | 1.06 (1.04,1.09) | 2.1E-06 | 3.7E-06 | 216 | 1.22 (1.11,1.35) | 6.6E-05 | 5.1E-04 | 1.15 (1.04, 1.27) | 6.8E-03 | 0.05 |
| QT_IND | 4615 | 1.06 (1.04,1.09) | 1.7E-06 | 3.3E-06 | 216 | 1.2 (1.08, 1.34) | 7.1E-04 | 2.7E-03 | 1.13 (1.01, 1.26) | 2.8E-02 | 0.13 |
| MC_MI | 4378 | 1.38 (1.18, 1.61) | 7.0E-05 | 1.1E-04 | 197 | 1.98 (1.07, 3.65) | 2.9E-02 | 0.05 | 1.44 (0.76, 2.71) | 0.26 | 0.73 |
| ECGBASENOVA | 4010 | 1.28 (1.2, 1.37) | 1.3E-13 | 3.7E-13 | 146 | 1.45 (1.04, 2.03) | 2.8E-02 | 0.05 | 1.13 (0.81, 1.59) | 0.47 | 0.81 |
| HDLC | 4624 | 0.86 (0.84,0.88) | 2.3E-35 | 2.6E-34 | 293 | 0.85 (0.77,0.94) | 1.3E-03 | 4.0E-03 | 0.99 (0.9, 1.1) | 0.88 | 1 |
| LDLC | 3838 | 1.04 (1.03,1.05) | 3.2E-18 | 1.8E-17 | 239 | 1.04 (1, 1.07) | 4.2E-02 | 0.07 | 1 (0.96, 1.03) | 0.92 | 1 |
| TG | 4284 | 1.41 (1.32, 1.5) | 4.3E-24 | 3.3E-23 | 259 | 1.31 (1, 1.72) | 0.05 | 0.08 | 0.93 (0.7, 1.23) | 0.63 | 0.85 |
| FG | 3577 | 1.88 (1.6, 2.2) | 5.3E-15 | 2.0E-14 | 227 | 2.68 (1.35, 5.33) | 4.8E-03 | 1.1E-02 | 1.43 (0.7, 2.9) | 0.32 | 0.73 |
| FI | 3196 | 1.27 (1.19, 1.35) | 5.1E-15 | 2.0E-14 | 180 | 1.26 (0.98, 1.62) | 0.08 | 0.12 | 0.99 (0.76, 1.28) | 0.94 | 1 |
| Creatinine | 2307 | 1.28 (1.03, 1.6) | 2.7E-02 | 3.3E-02 | 124 | 1.82 (0.7, 4.68) | 0.22 | 0.28 | 1.41 (0.53, 3.75) | 0.49 | 0.81 |
| CRP | 4383 | 1.13 (1.09, 1.16) | 7.3E-15 | 2.4E-14 | 293 | 1.27 (1.13, 1.44) | 9.5E-05 | 5.5E-04 | 1.13 (1, 1.28) | 0.06 | 0.23 |
| Meta-GRS | 2314 | 1.38 (1.32, 1.44) | 2.2E-47 | 5.1E-46 | 129 | 1.3 (1.08, 1.56) | 4.5E-03 | 1.1E-02 | 0.94 (0.78, 1.13) | 0.53 | 0.81 |

VE_N, Ventricular etopic beats, 0 vs 1; CV, Cornell Voltage (uV), per SD; CV_QRS, Cornell Voltage Production (uV)= (Cornell Voltage x QRS Duration / 1000), per SD; LVH_MINN, LVH by Minnesota Code, 0 vs 1; LVMI_Norm, Left Ventricular Mass Index (%), per SD; LVM_Norm, Left Ventricular Mass (g), per SD; PR_DUR, PR Interval (ms) , per SD; PROLO_PR, Prolonged PR interval, 0 vs 1; QRS_DUR, QRS Interval Duration (ms), per SD; QT_DUR, QT Interval Duration (ms), per SD; QTC_DUR, Bazett's Heart Rate-Corrected QT Interval (ms), per SD; QT_IND, Rautaharju's QT Prolongation Index (%), per SD; MC_MI, MI by Minnesota Code, 0 vs 1; ECGBASENOVA, ECG MI novacode, 0 vs 1; HDLC, high-density lipoprotein cholesterol, per 10 mg/dL of increase; LDLC, low-density lipoprotein cholesterol, per 10mg/dL of increase; TG, natural log transformed triglyceride; FG, natural log transformed fasting glucose; FI, natural log transformed fasting insulin; creatinine, natural log transformed; CRP, log-transformed C-reactive protein; wGRS, weighted genetic risk score, per standard deviation.

**Table S6. Annual estimated cumulative absolute risk over a 20 year follow up for subgroup of women with specific baseline characteristic related to age, smoking, lifestyle, and aspiring use using a joint multivariate Cox model.** We first estimate the absolute risk of a fatal presentation of incident CHD modeling the competing risk of a non-fatal CHD event followed by the absolute risk of a non-fatal CHD event. The absolute risks are then used to calculate the ratio of fatal CHD event over fatal and non-fatal CHD event.

|  | Yr1 | Yr2 | Yr3 | Yr4 | Yr5 | Yr6 | Yr7 | Yr8 | Yr9 | Yr10 | Yr11 | Yr12 | Yr13 | Yr14 | Yr15 | Yr16 | Yr17 | Yr18 | Yr19 | Yr20 |
| --- | --- | --- | --- | --- | --- | --- | --- | --- | --- | --- | --- | --- | --- | --- | --- | --- | --- | --- | --- | --- |
| Cumulative Probability of fatal incident CHD event (%) | | | | | | | | | | | | | | | | | | | | |
| age55+smk0+PA3 | 0.00 | 0.00 | 0.01 | 0.01 | 0.01 | 0.02 | 0.02 | 0.02 | 0.02 | 0.03 | 0.03 | 0.04 | 0.04 | 0.04 | 0.05 | 0.05 | 0.06 | 0.08 | 0.09 | 0.11 |
| age55+smk1+PA2 | 0.00 | 0.01 | 0.01 | 0.01 | 0.02 | 0.02 | 0.03 | 0.03 | 0.04 | 0.04 | 0.05 | 0.06 | 0.06 | 0.06 | 0.07 | 0.08 | 0.09 | 0.12 | 0.14 | 0.17 |
| age55+smk2+PA1 | 0.00 | 0.01 | 0.01 | 0.02 | 0.03 | 0.04 | 0.04 | 0.05 | 0.06 | 0.06 | 0.08 | 0.08 | 0.09 | 0.10 | 0.11 | 0.12 | 0.14 | 0.18 | 0.21 | 0.25 |
| age55+smk3+PA0 | 0.01 | 0.01 | 0.02 | 0.03 | 0.04 | 0.05 | 0.07 | 0.08 | 0.09 | 0.10 | 0.12 | 0.13 | 0.14 | 0.15 | 0.16 | 0.18 | 0.21 | 0.28 | 0.32 | 0.39 |
| age65+smk0+PA3 | 0.01 | 0.01 | 0.02 | 0.03 | 0.04 | 0.05 | 0.06 | 0.07 | 0.08 | 0.09 | 0.11 | 0.12 | 0.13 | 0.14 | 0.16 | 0.18 | 0.20 | 0.26 | 0.31 | 0.37 |
| age65+smk1+PA2 | 0.01 | 0.02 | 0.03 | 0.05 | 0.06 | 0.08 | 0.10 | 0.11 | 0.13 | 0.14 | 0.17 | 0.19 | 0.20 | 0.22 | 0.24 | 0.27 | 0.31 | 0.40 | 0.47 | 0.56 |
| age65+smk2+PA1 | 0.01 | 0.03 | 0.05 | 0.07 | 0.10 | 0.12 | 0.15 | 0.17 | 0.20 | 0.22 | 0.26 | 0.28 | 0.30 | 0.33 | 0.36 | 0.41 | 0.47 | 0.61 | 0.71 | 0.85 |
| age65+smk3+PA0 | 0.02 | 0.05 | 0.08 | 0.11 | 0.15 | 0.18 | 0.23 | 0.26 | 0.30 | 0.33 | 0.39 | 0.43 | 0.46 | 0.50 | 0.55 | 0.61 | 0.71 | 0.92 | 1.07 | 1.28 |
| age75+smk0+PA3 | 0.02 | 0.05 | 0.07 | 0.10 | 0.14 | 0.18 | 0.22 | 0.25 | 0.29 | 0.32 | 0.37 | 0.41 | 0.44 | 0.48 | 0.53 | 0.59 | 0.68 | 0.88 | 1.03 | 1.23 |
| age75+smk1+PA2 | 0.03 | 0.07 | 0.11 | 0.15 | 0.21 | 0.27 | 0.33 | 0.38 | 0.43 | 0.48 | 0.56 | 0.62 | 0.67 | 0.73 | 0.80 | 0.89 | 1.02 | 1.32 | 1.55 | 1.85 |
| age75+smk2+PA1 | 0.05 | 0.11 | 0.17 | 0.24 | 0.32 | 0.41 | 0.51 | 0.58 | 0.66 | 0.73 | 0.86 | 0.95 | 1.01 | 1.10 | 1.21 | 1.34 | 1.54 | 1.99 | 2.33 | 2.77 |
| age75+smk3+PA0 | 0.08 | 0.17 | 0.26 | 0.36 | 0.49 | 0.62 | 0.77 | 0.88 | 1.00 | 1.11 | 1.29 | 1.43 | 1.53 | 1.66 | 1.82 | 2.02 | 2.31 | 2.99 | 3.49 | 4.13 |
| age75+smk0+PA3+ASA | 0.02 | 0.04 | 0.06 | 0.08 | 0.11 | 0.14 | 0.18 | 0.20 | 0.23 | 0.26 | 0.30 | 0.33 | 0.36 | 0.39 | 0.43 | 0.48 | 0.55 | 0.71 | 0.83 | 0.99 |
| age75+smk1+PA2+ASA | 0.03 | 0.06 | 0.09 | 0.13 | 0.17 | 0.22 | 0.27 | 0.31 | 0.35 | 0.39 | 0.46 | 0.51 | 0.54 | 0.59 | 0.65 | 0.72 | 0.83 | 1.07 | 1.26 | 1.49 |
| age75+smk2+PA1+ASA | 0.04 | 0.09 | 0.14 | 0.19 | 0.26 | 0.33 | 0.41 | 0.47 | 0.54 | 0.60 | 0.70 | 0.77 | 0.82 | 0.89 | 0.98 | 1.09 | 1.25 | 1.61 | 1.88 | 2.23 |
| age75+smk3+PA0+ASA | 0.06 | 0.14 | 0.21 | 0.29 | 0.40 | 0.50 | 0.63 | 0.72 | 0.81 | 0.90 | 1.05 | 1.16 | 1.24 | 1.35 | 1.48 | 1.64 | 1.87 | 2.41 | 2.81 | 3.33 |
| Cumulative Probability of non-fatal incident CHD event (%) | | | | | | | | | | | | | | | | | | | | |
| age55+smk0+PA3 | 0.18 | 0.37 | 0.57 | 0.77 | 0.99 | 1.20 | 1.41 | 1.61 | 1.81 | 1.99 | 2.17 | 2.34 | 2.48 | 2.60 | 2.69 | 2.79 | 2.94 | 3.14 | 3.35 | 3.66 |
| age55+smk1+PA2 | 0.21 | 0.44 | 0.67 | 0.91 | 1.17 | 1.41 | 1.66 | 1.89 | 2.13 | 2.35 | 2.56 | 2.76 | 2.92 | 3.06 | 3.18 | 3.29 | 3.47 | 3.70 | 3.94 | 4.31 |
| age55+smk2+PA1 | 0.25 | 0.52 | 0.79 | 1.08 | 1.38 | 1.67 | 1.96 | 2.24 | 2.51 | 2.77 | 3.02 | 3.25 | 3.45 | 3.61 | 3.74 | 3.88 | 4.08 | 4.35 | 4.64 | 5.07 |
| age55+smk3+PA0 | 0.30 | 0.61 | 0.93 | 1.27 | 1.62 | 1.97 | 2.32 | 2.64 | 2.96 | 3.27 | 3.56 | 3.83 | 4.06 | 4.25 | 4.41 | 4.56 | 4.81 | 5.12 | 5.46 | 5.97 |
| age65+smk0+PA3 | 0.29 | 0.59 | 0.91 | 1.23 | 1.58 | 1.91 | 2.25 | 2.56 | 2.88 | 3.17 | 3.46 | 3.72 | 3.94 | 4.12 | 4.28 | 4.43 | 4.67 | 4.97 | 5.30 | 5.79 |
| age65+smk1+PA2 | 0.34 | 0.70 | 1.07 | 1.46 | 1.86 | 2.26 | 2.65 | 3.02 | 3.39 | 3.73 | 4.07 | 4.38 | 4.64 | 4.85 | 5.03 | 5.21 | 5.49 | 5.85 | 6.23 | 6.81 |
| age65+smk2+PA1 | 0.40 | 0.82 | 1.26 | 1.72 | 2.19 | 2.66 | 3.13 | 3.55 | 3.99 | 4.40 | 4.79 | 5.15 | 5.46 | 5.71 | 5.92 | 6.13 | 6.45 | 6.87 | 7.32 | 7.99 |
| age65+smk3+PA0 | 0.48 | 0.97 | 1.49 | 2.03 | 2.59 | 3.14 | 3.68 | 4.19 | 4.70 | 5.17 | 5.64 | 6.06 | 6.42 | 6.71 | 6.95 | 7.20 | 7.57 | 8.06 | 8.59 | 9.36 |
| age75+smk0+PA3 | 0.46 | 0.94 | 1.45 | 1.97 | 2.51 | 3.04 | 3.57 | 4.06 | 4.56 | 5.02 | 5.47 | 5.88 | 6.23 | 6.51 | 6.75 | 6.99 | 7.36 | 7.83 | 8.34 | 9.09 |
| age75+smk1+PA2 | 0.55 | 1.12 | 1.71 | 2.32 | 2.96 | 3.59 | 4.21 | 4.78 | 5.37 | 5.91 | 6.43 | 6.91 | 7.31 | 7.64 | 7.92 | 8.20 | 8.63 | 9.18 | 9.77 | 10.64 |
| age75+smk2+PA1 | 0.65 | 1.32 | 2.02 | 2.74 | 3.49 | 4.22 | 4.95 | 5.63 | 6.31 | 6.94 | 7.55 | 8.11 | 8.58 | 8.96 | 9.28 | 9.60 | 10.10 | 10.73 | 11.41 | 12.41 |
| age75+smk3+PA0 | 0.77 | 1.56 | 2.38 | 3.23 | 4.11 | 4.97 | 5.83 | 6.61 | 7.41 | 8.14 | 8.85 | 9.49 | 10.04 | 10.48 | 10.85 | 11.22 | 11.79 | 12.52 | 13.30 | 14.44 |
| age75+smk0+PA3+ASA | 0.55 | 1.12 | 1.72 | 2.33 | 2.98 | 3.60 | 4.23 | 4.81 | 5.40 | 5.94 | 6.47 | 6.95 | 7.36 | 7.69 | 7.97 | 8.25 | 8.68 | 9.23 | 9.83 | 10.71 |
| age75+smk1+PA2+ASA | 0.65 | 1.32 | 2.03 | 2.75 | 3.51 | 4.25 | 4.98 | 5.66 | 6.35 | 6.98 | 7.60 | 8.16 | 8.63 | 9.02 | 9.34 | 9.67 | 10.17 | 10.81 | 11.50 | 12.52 |
| age75+smk2+PA1+ASA | 0.77 | 1.56 | 2.39 | 3.24 | 4.13 | 5.00 | 5.86 | 6.65 | 7.46 | 8.19 | 8.91 | 9.56 | 10.11 | 10.56 | 10.94 | 11.31 | 11.89 | 12.63 | 13.42 | 14.59 |
| age75+smk3+PA0+ASA | 0.91 | 1.85 | 2.82 | 3.82 | 4.87 | 5.88 | 6.89 | 7.81 | 8.74 | 9.60 | 10.43 | 11.19 | 11.82 | 12.34 | 12.77 | 13.20 | 13.86 | 14.71 | 15.62 | 16.95 |
| Ratio of the cumulative probability of an incident fatal CHD event / (cumulative probability of all incident fatal + non-fatal CHD events) (%) | | | | | | | | | | | | | | | | | | | | |
| age55+smk0+PA3 | 1.02 | 1.11 | 1.10 | 1.12 | 1.21 | 1.26 | 1.34 | 1.35 | 1.36 | 1.38 | 1.48 | 1.53 | 1.55 | 1.61 | 1.71 | 1.84 | 2.00 | 2.44 | 2.68 | 2.93 |
| age55+smk1+PA2 | 1.32 | 1.43 | 1.42 | 1.45 | 1.56 | 1.62 | 1.72 | 1.74 | 1.76 | 1.78 | 1.91 | 1.96 | 1.99 | 2.07 | 2.19 | 2.36 | 2.57 | 3.13 | 3.44 | 3.75 |
| age55+smk2+PA1 | 1.70 | 1.85 | 1.83 | 1.86 | 2.01 | 2.08 | 2.21 | 2.23 | 2.26 | 2.29 | 2.45 | 2.52 | 2.56 | 2.66 | 2.81 | 3.03 | 3.29 | 4.00 | 4.39 | 4.78 |
| age55+smk3+PA0 | 2.19 | 2.37 | 2.35 | 2.40 | 2.58 | 2.67 | 2.84 | 2.87 | 2.90 | 2.93 | 3.14 | 3.23 | 3.28 | 3.41 | 3.60 | 3.87 | 4.21 | 5.10 | 5.60 | 6.09 |
| age65+smk0+PA3 | 2.15 | 2.34 | 2.32 | 2.36 | 2.54 | 2.63 | 2.80 | 2.83 | 2.86 | 2.89 | 3.10 | 3.19 | 3.23 | 3.36 | 3.55 | 3.82 | 4.15 | 5.03 | 5.52 | 6.00 |
| age65+smk1+PA2 | 2.77 | 3.00 | 2.98 | 3.03 | 3.26 | 3.38 | 3.59 | 3.62 | 3.66 | 3.71 | 3.97 | 4.08 | 4.14 | 4.30 | 4.54 | 4.88 | 5.30 | 6.40 | 7.01 | 7.61 |
| age65+smk2+PA1 | 3.55 | 3.85 | 3.81 | 3.89 | 4.17 | 4.32 | 4.59 | 4.63 | 4.68 | 4.74 | 5.06 | 5.21 | 5.28 | 5.48 | 5.79 | 6.21 | 6.73 | 8.10 | 8.85 | 9.60 |
| age65+smk3+PA0 | 4.54 | 4.92 | 4.88 | 4.97 | 5.33 | 5.52 | 5.85 | 5.90 | 5.97 | 6.04 | 6.45 | 6.63 | 6.72 | 6.97 | 7.35 | 7.87 | 8.52 | 10.20 | 11.12 | 12.03 |
| age75+smk0+PA3 | 4.48 | 4.85 | 4.81 | 4.90 | 5.25 | 5.44 | 5.77 | 5.82 | 5.89 | 5.96 | 6.36 | 6.54 | 6.63 | 6.88 | 7.25 | 7.76 | 8.41 | 10.07 | 10.98 | 11.88 |
| age75+smk1+PA2 | 5.71 | 6.18 | 6.13 | 6.24 | 6.69 | 6.92 | 7.34 | 7.40 | 7.48 | 7.56 | 8.06 | 8.29 | 8.40 | 8.70 | 9.17 | 9.80 | 10.58 | 12.61 | 13.71 | 14.79 |
| age75+smk2+PA1 | 7.26 | 7.85 | 7.78 | 7.92 | 8.47 | 8.76 | 9.28 | 9.36 | 9.45 | 9.56 | 10.18 | 10.45 | 10.58 | 10.96 | 11.52 | 12.28 | 13.23 | 15.66 | 16.96 | 18.24 |
| age75+smk3+PA0 | 9.19 | 9.92 | 9.83 | 10.00 | 10.69 | 11.04 | 11.67 | 11.77 | 11.88 | 12.02 | 12.76 | 13.09 | 13.25 | 13.70 | 14.38 | 15.29 | 16.41 | 19.26 | 20.78 | 22.25 |
| age75+smk0+PA3+ASA | 3.13 | 3.39 | 3.36 | 3.43 | 3.68 | 3.81 | 4.05 | 4.09 | 4.13 | 4.18 | 4.47 | 4.60 | 4.66 | 4.84 | 5.11 | 5.48 | 5.94 | 7.15 | 7.82 | 8.48 |
| age75+smk1+PA2+ASA | 4.01 | 4.34 | 4.30 | 4.38 | 4.70 | 4.87 | 5.17 | 5.22 | 5.27 | 5.34 | 5.70 | 5.86 | 5.94 | 6.16 | 6.50 | 6.95 | 7.53 | 9.02 | 9.84 | 10.65 |
| age75+smk2+PA1+ASA | 5.12 | 5.54 | 5.49 | 5.60 | 6.00 | 6.21 | 6.58 | 6.64 | 6.71 | 6.79 | 7.24 | 7.43 | 7.53 | 7.81 | 8.22 | 8.78 | 9.49 | 11.31 | 12.30 | 13.28 |
| age75+smk3+PA0+ASA | 6.52 | 7.05 | 6.99 | 7.12 | 7.62 | 7.88 | 8.34 | 8.41 | 8.50 | 8.60 | 9.15 | 9.39 | 9.51 | 9.85 | 10.36 | 11.04 | 11.89 | 14.08 | 15.26 | 16.42 |

Absolute risk is calculated for women aged 55 (age55), 65 (age65), and 75 (age75) at baseline with and without healthy lifestyles as defined by degree of smoking and weekly physical activity. The level of physical activity was categorized into four groups: sedentary (PA0): ≤100 MET minutes/week (min/wk); low (PA1): >100 and ≤500 MET min/wk; moderate (PA2): >500 and ≤1200 MET min/wk; and high (PA3): >1200 MET min/wk. The level of smoking was categorized into five groups: <1 (smk0), 1-4 (smk1), 5-34 (smk2), 35-44 (smk3), ≥ 45 cigarettes/day (not modelled here). For women aged 75 years, absolute risk is also calculated with and without use of aspirin (ASA).
